# Genome-wide association study of problematic opioid prescription use in 132,113 23andMe research participants of European ancestry

**DOI:** 10.1101/2021.06.03.21258260

**Authors:** Sandra Sanchez-Roige, Pierre Fontanillas, Mariela V Jennings, Sevim Bianchi, Yuye Huang, Alex Hatoum, Julia Sealock, Lea K Davis, Sarah L Elson, 23andMe Research Team, Abraham A Palmer

## Abstract

The growing prevalence of opioid use disorder (**OUD**) constitutes an urgent health crisis. Ample evidence indicates that risk for OUD is heritable. As a surrogate (or proxy) for OUD, we explored the genetic basis of using prescription opioids ‘not as prescribed’. We hypothesized that misuse of opiates might be a heritable risk factor for OUD. To test this hypothesis, we performed a genome-wide association study (**GWAS**) of problematic opioid use (**POU**) in 23andMe research participants of European ancestry (N=132,113; 21% cases). We identified two genome-wide significant loci (rs3791033, an intronic variant of *KDM4A*; rs640561, an intergenic variant near *LRRIQ3*). POU showed a positive genetic correlation with the largest available GWAS of opioid dependence and OUD (rg=0.64-0.80). We also identified numerous additional genetic correlations with POU, including alcohol dependence (rg=0.74), smoking initiation (rg=0.63), pain relief medication intake (rg=0.49), major depressive disorder (rg=0.44), chronic pain (rg=0.42), insomnia (rg=0.39), and loneliness (rg=0.28). Although POU was positively genetically correlated with risk-taking (rg=0.38), conditioning POU on risk-taking did not substantially alter the magnitude or direction of these genetic correlations, suggesting that POU does not simply reflect a genetic tendency towards risky behavior. Lastly, we performed phenome- and lab-wide association analyses, which uncovered additional phenotypes that were associated with POU, including respiratory failure, insomnia, ischemic heart disease, and metabolic and blood-related biomarkers. We conclude that opioid misuse can be measured in population-based cohorts and provides a cost-effective complementary strategy for understanding the genetic basis of OUD.

## INTRODUCTION

Opioid use disorders (**OUD**) represent a global epidemic [1]. Every day, 128 people in the United States die after overdosing on opiates. The pathway to opiate addiction has changed over the last few decades. In the 1960s, more than 80% of people who began using opioids initiated with heroin [2]; by 2013, nearly 80% of opioid users reported that their first regular opioid was a prescription pain reliever [3]. The misuse of and addiction to opiates—including prescription pain relievers, heroin, and synthetic opioids such as fentanyl— is thus a serious emergency that affects public health as well as social and economic welfare. More recently, the COVID-19 pandemic made it increasingly difficult for individuals with OUD to access treatment and impacted mental health, triggering both initial and continued use of opioids [4], which is likely to further increase rates of OUD [5].

Although OUD is known to be moderately heritable [6, 7], genomic discovery has been severely limited due to the complexity of obtaining large, well-characterized samples of cases and opiate-exposed controls [6, 8–14]. The largest genome-wide association study (**GWAS**) to date (∼10K cases) identified only one locus in *OPRM1*, which encodes the µ-opioid receptor [7]. Larger sample sizes are urgently needed to identify additional loci associated with OUD.

An alternative, complementary approach is to study the genetic liability for OUD across different stages, particularly the transition from use to misuse. By taking this approach, we can explore specific aspects of OUD liability, such as initial use, subjective drug response, transition to hazardous use, dependence, withdrawal and relapse. This strategy can be applied in population-based cohorts, allowing for a dramatic increase in sample sizes for a fraction of the cost.

In the present study, we pursued a novel strategy in which we measured problematic prescription opioid use (**POU**) in 23andMe research participants (n=132,113) of European ancestry. We asked a single question “Have you ever in your life used prescription painkillers (taken not as prescribed), e.g., Vicodin, Oxycontin?”, and conducted a GWAS defining cases as those who answered ‘yes’. We used a variety of bioinformatic analyses to explore the relationship between POU and OUD as well as other substance use disorder related behaviors. We also explored POU’s genetic relationship with various psychiatric, behavioral and medical conditions.

## MATERIALS AND METHODS

### GWAS cohort and phenotype

We utilized a cohort of 132,113 male and female research participants of European ancestry. All participants were drawn from the customer base of 23andMe, Inc., a direct-to-consumer genetics company. Participants provided informed consent and participated in the research online, under a protocol approved by the external AAHRPP-accredited IRB, Ethical & Independent Review Services (www.eandireview.com). During four months in 2015 and 14 months in 2018-2020, participants responded to a decision-making survey that, depending on branching logic, included up to 139 questions pertaining to aspects of impulsivity and substance use and abuse. A single item in this survey asked, “Have you ever in your life used prescription painkillers (taken not as prescribed), e.g., Vicodin, Oxycontin?”. A total of 132,113 individuals responded, with 27,805 (10,164 males) answering ‘yes’ (cases) and the remaining 104,308 (36,246 males) answering ‘no’ (controls). Controls were not screened for prior opioid use, so they represent a combination of individuals who have taken opioids but only as prescribed, and others who have never taken an opioid. Only individuals who were categorized as being of European ancestry based on empirical genotype data [15] were included in this study. Demographic information about this sample is presented in **Supplementary Table 1**.

### Genome-wide association analysis

DNA extraction and genotyping were performed on saliva samples by CLIA-certified and CAP-accredited clinical laboratories of Laboratory Corporation of America. Quality control, imputation, and genome-wide analysis were performed by 23andMe (**Supplementary Table 2**; see **Supplemental Material** and [16, 17] for further information about genotyping, imputation and quality control).

As previously described [17, 18], 23andMe’s analysis pipeline performs logistic regression assuming an additive model for allelic effects (**Supplementary Material**). Covariates included age (inverse-normal transformed), sex, the top five principal components of ancestry, and indicator variables for genotyping platforms. P-values were not corrected for genomic control.

### Biological annotation, gene and transcriptome-based association analyses

We used a variety of bioinformatic methods to further characterize the loci identified by the GWAS. First, we used the default version (v1.3.6a) of the FUMA web-based platform [19] to identify independent SNPs (r^2^<0.10) and to study their functional consequences. We also used MAGMA v1.08 [19, 20] to perform competitive gene-set and pathway analyses. SNPs were mapped to 18,546 protein-coding genes from Ensembl (build 85). We applied a Bonferroni correction based on the total number of genes tested (*p*<2.56E-06). Gene-sets were obtained from Msigdb v7.0 (“Curated gene sets”, “GO terms”). We also used Hi-C coupled MAGMA (**H-MAGMA**; [21]) to assign non-coding (intergenic and intronic) SNPs to genes based on their chromatin interactions. Exonic and promoter SNPs were assigned to genes based on physical position. H-MAGMA uses four Hi-C datasets, which were derived from fetal brain, adult brain, iPSC-derived neurons and iPSC-derived astrocytes (https://github.com/thewonlab/H-MAGMA). We applied a Bonferroni correction based on the total number of gene-tissue pairs tested (*p*<9.55E-07).

Lastly, we used S-MultiXcan v0.7.0 (an extension of S-PrediXcan v0.6.2 [22]) to identify specific eQTL-linked genes associated with POU. This approach uses genetic information to predict transcript abundance in 13 brain tissues, and tests whether the predicted transcripts correlate with POU. S-PrediXcan uses pre-computed tissue weights from the Genotype-Tissue Expression (**GTEx**) v8 project database (https://www.gtexportal.org/) as the reference transcriptome dataset. For S-PrediXcan and S-MultiXcan analyses we chose to use sparse (elastic net) prediction models, which are available at http://predictdb.hakyimlab.org/. We applied a conservative Bonferroni correction based on the total number of gene-tissue pairs tested (14,159 gene-tissue pairs tested; *p*<3.53E-06).

### Gene-drug interaction analysis

We examined the 17 genes that were significantly associated with POU in the MAGMA gene-based analysis (10% FDR) for known interactions with prescription medications using the Drug Gene Interaction Database v3.0 (dgidb.genome.wustl.edu) [23]. We used the Anatomical Therapeutic Chemical (**ATC**) classification system to determine the second level classification of each medication we identified. ATC classifications for medications were retrieved from the Kyoto Encyclopedia of Genes and Genomics (**KEGG**; https://www.genome.jp/kegg/drug/) and the World Health Organization Collaborating Center for Drug Statistics Methodology (https://www.whocc.no/atc_ddd_index/). We used the R package *circlize* v0.4.1 [24] to visualize the interactions between each gene and the ATC classifications of drugs it interacts with.

### Heritability

We used the LD Score regression (**LDSC** [25]) python package to estimate the heritability explained by SNPs (SNP-h_2_). We used pre-computed LD scores (“eur_w_ld_chr/”), which are publicly available (https://data.broadinstitute.org/alkesgroup/LDSCORE/). LD scores were computed for every SNP using individuals from European ancestry from the 1000 Genomes Project. We restricted the analysis to well-imputed SNPs, filtered to HapMap3 SNPs, with MAF above 1%. We removed InDels, structural variants, strand-ambiguous SNPs and SNPs with extremely large effect sizes (χ^2^>80). Heritability was calculated on the liability scale by accounting for differences in population prevalence (4%) and sample prevalence (21%). The population prevalence was retrieved from the 2018 National Survey on Drug Use and Health [26].

### Genetic correlation analyses

We used LDhub or local LDSC [25] to calculate genetic correlations (*r*_*g*_) between POU and 935 other traits or diseases (852 from LDHub and 83 local) [25]. Local traits were selected based on previously known phenotypic associations between OUD and other substance use disorder phenotypes and related traits (e.g., cannabis use disorder, various measures of impulsivity) not available on LDhub. We used the standard Benjamini–Hochberg false discovery rate correction (FDR 5%) to correct for multiple testing. We also calculated a Bonferroni correction for 935 comparisons (*p*<5.35E-05); however, this correction is overly conservative because many of the 935 traits are highly correlated with one another.

### mtCOJO

We used mtCOJO [24] to individually condition the POU summary statistics on loci associated with other comorbid traits, including risk-taking behavior [27], smoking [28], cannabis use disorder [29], alcohol dependence [30], chronic pain [31], and OUD [32]. This analysis allowed us to examine whether the genetic associations with POU would be preserved when controlling for those covariate phenotypes. To test as many SNPs while preserving computational efficiency, we used a *p-*value threshold of 2.00E-05, 5.00E-08,1.00E-05, 1.00E-05, 5.00E-08, and 1.00E-05, respectively for risk-taking behavior, smoking, cannabis use disorder, alcohol dependence, chronic pain, and OUD. We then computed genetic correlations using the POU summary statistics adjusted for the covariates of interest.

### Unsupervised learning to determine POU clustering

Previous studies have shown that consumption and misuse/dependence phenotypes have a distinct genetic architecture. To explore whether POU clustered more with consumption or misuse/dependence phenotypes we used a data-driven unsupervised machine learning method known as agglomerative hierarchical clustering analysis (**HCA**) [33, 34]. HCA forms clusters iteratively by creating groups and successively joining or splitting those groups based on a prespecified algorithm [33]. Agglomerative nesting (**AGNES**) is a bottom-up process focused on individual traits to structure. Agglomerative clustering was chosen as this allowed us to compare different algorithms to maximize for the dissimilarity on each branch, with Ward’s minimum variance method performing best. All models were fit in R using the *cluster* package [33].

The product of HCA is a dendrogram, formed with multiple brackets called “branches”. Phenotypes on the same branch are more similar to each other based on their pairwise genetic associations with each other and with all other phenotypes on that branch. Branches can form subbranches of more specific clustering.

### Polygenic risk score phenome-wide and lab-wide association scans in BioVU

To explore whether pleiotropic effects for POU are associated with an array of other medical ailments and biomarkers, we performed phenome-wide and lab-wide association scans (PheWAS, LabWAS), respectively. These analyses were conducted using data from the Vanderbilt University Medical Center (**VUMC**). The project was approved by the VUMC Institutional Review Board (IRB #160302, #172020, #190418). VUMC is an integrated health system with individual-level health data from electronic health records (**EHR**) for about 3.2 million patients. The VUMC biobank contains clinical data from EHR as well as biomarkers obtained from laboratory assessments. A portion of the individuals from VUMC also have accompanying array genotyping data. This cohort, with over 66,903 patients, is called BioVU [35, 36].

For each of the unrelated genotyped individuals of European ancestry from BioVU, we computed polygenic risk scores (**PRS**) for POU using the PRS-CS “auto” version [35]. Genotyping and quality control for this cohort have been extensively described [36, 37].

To identify associations between the PRS for POU and clinical phenotypes, we performed a PheWAS in BioVU. We fitted a logistic regression model to each of 1,338 case/control disease phenotypes (“phecodes”) to estimate the odds of each diagnosis given the POU PRS, while adjusting for sex, median age of the longitudinal EHR, and the first 10 PCs. Analyses were conducted using the PheWAS v0.12 R package [38]. We required the presence of at least two International Disease Classification codes mapped to a PheWAS disease category (Phecode Map 1.2; https://phewascatalog.org/phecodes) and a minimum of 100 cases for inclusion of a phecode. The disease phenotypes included 145 circulatory system, 120 genitourinary, 119 endocrine/metabolic, 125 digestive, 117 neoplasms, 91 musculoskeletal, 85 sense organs, 76 injuries & poisonings, 65 dermatological, 76 respiratory, 69 neurological, 64 mental disorders, 42 infectious diseases, 42 hematopoietic, 34 congenital anomalies, 37 symptoms, and 31 pregnancy complications.

To identify associations between POU PRS and biomarkers, we performed a LabWAS [37] in BioVU. We implemented the pipeline already established by Dennis et al [37]. Broadly, LabWAS uses the median, INT-transformed age-adjusted values from the QualityLab pipeline in a linear regression to determine the association with the input POU PRS variable. We controlled for the same covariates as for the PheWAS analyses, excluding median age because the pipeline corrects for age using cubic splines with 4 knots.

Lastly, to explore whether the POU PRS effects could be mediated by the diagnosis of substance use disorders (**SUD**) or OUD in EHR, we repeated the PheWAS and LabWAS analyses after adjustment for SUD (N=2,304 cases) and OUD (N=1,799 cases). We applied FDR (5%) correction for both PheWAS and LabWAS analyses.

## RESULTS

### Genome-wide association analysis, biological annotation, gene and transcriptome-based association analyses

We examined 11,311,983 SNPs in all 132,113 study participants. The inflation factor of the GWAS was λ_GC_=1.097 with an LDSC intercept of 1.004 (SE=0.008), suggesting that the majority of the inflation was due to polygenicity. The SNP heritability of POU on the liability scale was SNP-h_2_=0.04 +/- 0.01.

We identified two genome-wide significant loci: rs3791033 near the genes *KDM4A* and *PTPRF* (*p*=3.80E-08), and rs640561 near *LRRIQ3* (*p*=3.80E-08; **Figure 1, Supplementary Table 3** and **Supplementary Figures 1-2** for locus zoom plots). Both loci are on chromosome 1, but rs3791033 and rs640561 are independent (r^2^<0.001).

**Figure 1.**
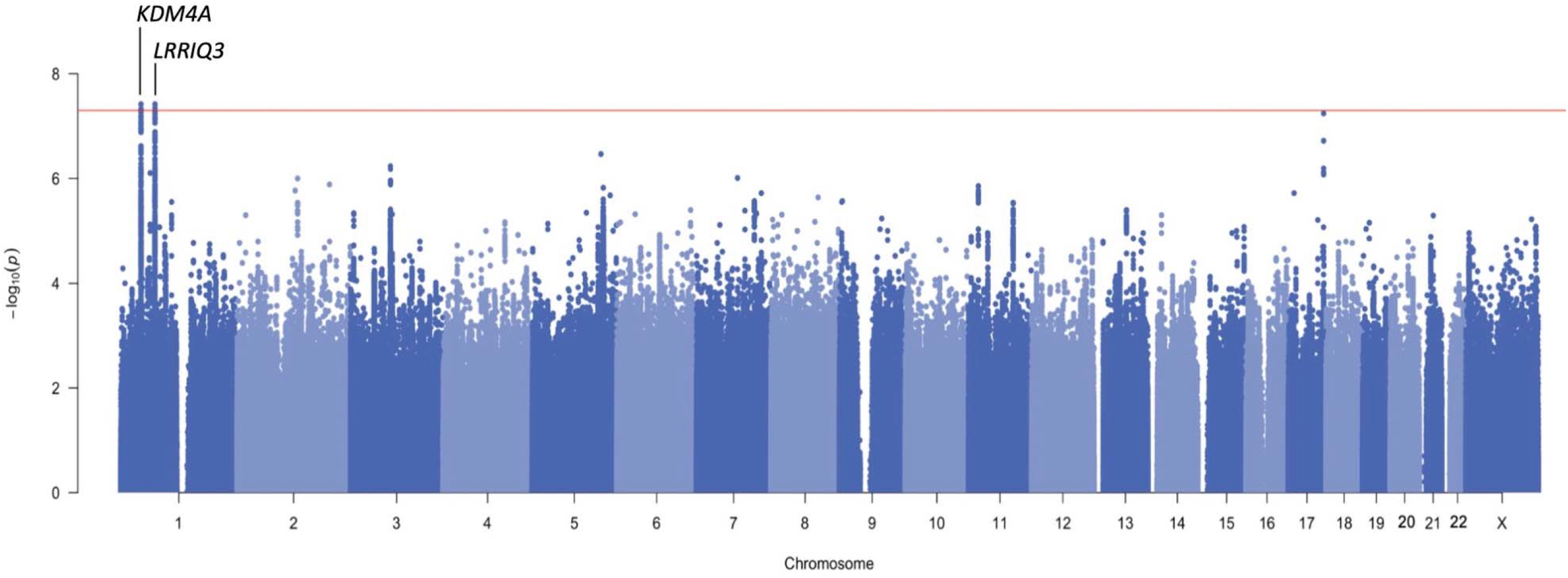
Genome-wide association analysis for problematic opioid use. The nearest genes are labeled for the 2 lead SNPs (rs3791033, rs640561). The x-axis shows chromosomal position and the y-axis shows significance on –log10 scale. The horizontal red line denotes genome-wide significance (*p*=5.00E-08).

**Figure 2.**
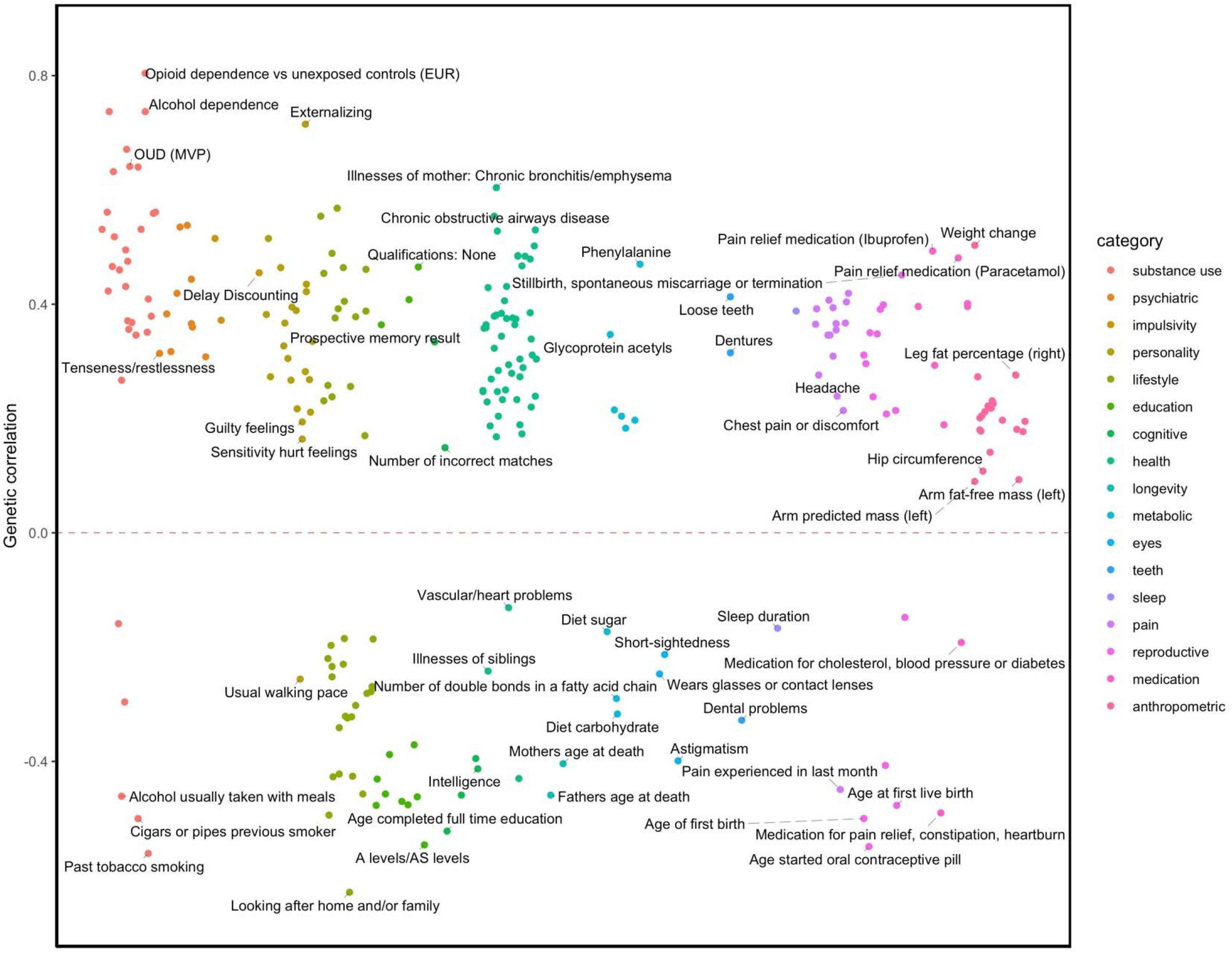
LDSC FDR-significant genetic correlations with POU. Traits with positive genetic correlation (*r*_*g*_) values are plotted above the line; traits with negative *r*_*g*_ values are plotted below the line. All traits surpass 5% FDR correction for multiple testing.

A phenome-wide scan in the UK Biobank (UKB; N>360,000) revealed that rs3791033 has been previously implicated in smoking phenotypes (e.g., ever smoked regularly, *p*=1.01E-12), other psychiatric conditions, such as ADHD (*p*=2.76E-10), and educational outcomes, such as educational attainment (*p*=1.01E-10). *KDM4A*, which is the nearest gene, has been previously implicated with similar traits across several independent GWAS studies (*e*.*g*., [27, 39, 40]; **Supplementary Table 4**). The other gene in this region, *PTPRF*, has been previously identified in studies of smoking and other substance use behaviors [18, 27, 41]; **Supplementary Table 4**).

For the second genome-wide significant SNP (rs640561), a phenome-wide scan in UKB revealed nominal associations (*p*>7.00E-03) with neurological, metabolic and other psychiatric traits, such as ADHD (**Supplementary Table 4**). *LRRIQ3*, which is the nearest gene, has previously been associated with lifetime smoking [27].

We did not observe any GWAS-significant (*p*=5.00E-08) or even nominally significant (*p*<0.05) associations with any of the SNPs that have been previously associated with OUD and opioid dependence ([6, 8–13]; **Supplementary Table 5**). However, the largest available GWAS of OUD supported our finding for rs640561 (*p*=6.42E-03; **Supplementary Table 5**).

In addition to the GWAS, we performed several gene-based analyses. *KDM4A* was implicated by both MAGMA (FDR 5%) and H-MAGMA (**Supplementary Tables 6, 7**). *PTPRF* was implicated by H-MAGMA (**Supplementary Table 7**). None of these analyses identified *LRRIQ3*. H-MAGMA analysis also identified *ARTN* (previously showing a nominal association with tea consumption in UKB; **Supplementary Table 7**). S-MultiXcan did not identify any significant genes (**Supplementary Table 8**).

Lastly, gene-set analysis in MAGMA identified one significant gene set, which is involved in the activation of Phospholipase D (**Supplementary Table 9**). Intriguingly, activation of Phospholipase D2 modulates agonist-induced µ-opioid receptor desensitization and resensitization [42].

### Gene-Drug Analysis

For this analysis we relaxed the FDR threshold from 5 to 10%, which produced a set of 17 genes; 3 of these 17 genes (*KDM4A, PTPRF, CACNA2D2*) had a total of 464 interactions with drugs belonging to ATC drug classes (see **Figure 4** and **Supplementary Table 10**). The most abundant second-level ATC classifications identified were antibacterials for systemic use, psycholeptics, and gynecological antiinfectives and antiseptics. Interactions with *KDM4A* included both dopamine agonists and agents, as well as adrenergic agents, drugs used in alcohol dependence such as disulfiram, opioid anaesthetics, such as sufentanil citrate, and antidepressants.

**Figure 3.**
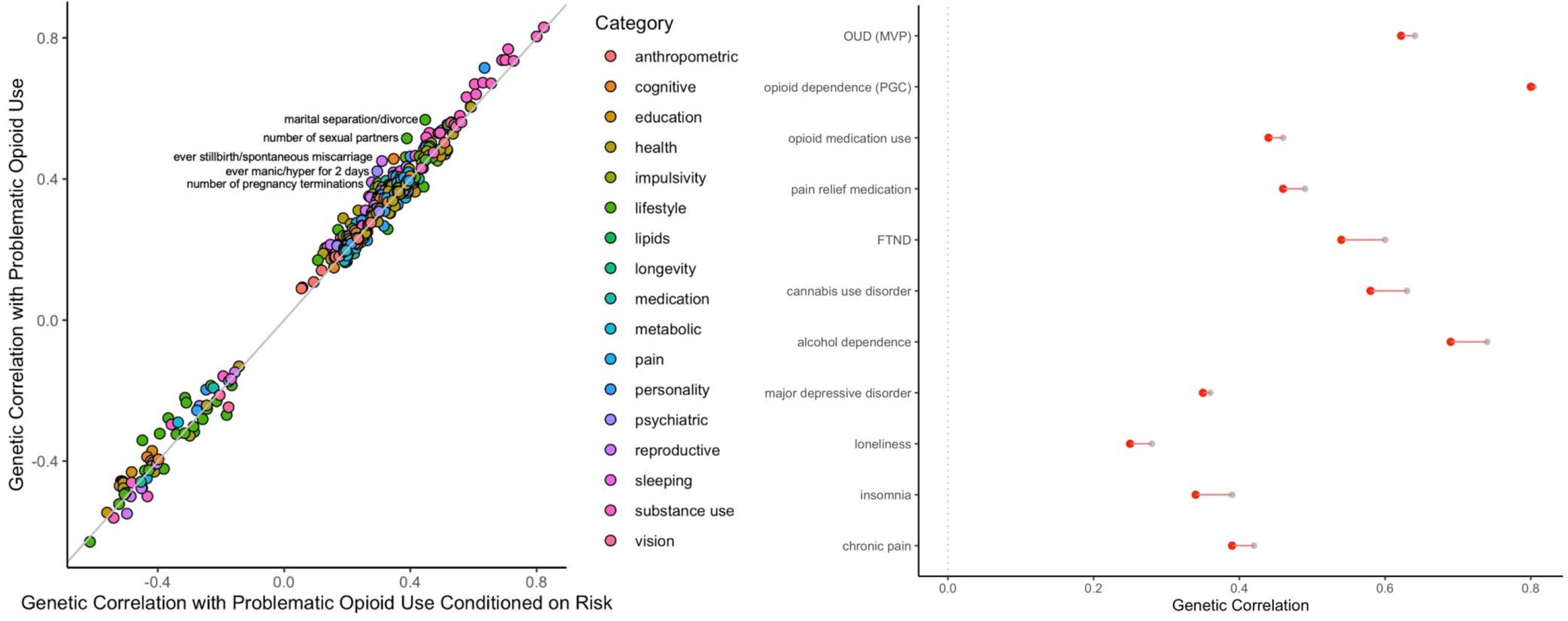
Genetic correlations (*r*_*g*_) with POU before and after conditioning on risk-taking behavior. All FDR-significant results are plotted on the left panel, selected relevant traits are shown on the right panel (original *r*_*g*_ in grey; corrected *r*_*g*_ in red). The top 5 traits with biggest change in *r*_*g*_ value are labelled (left).

**Figure 4.**
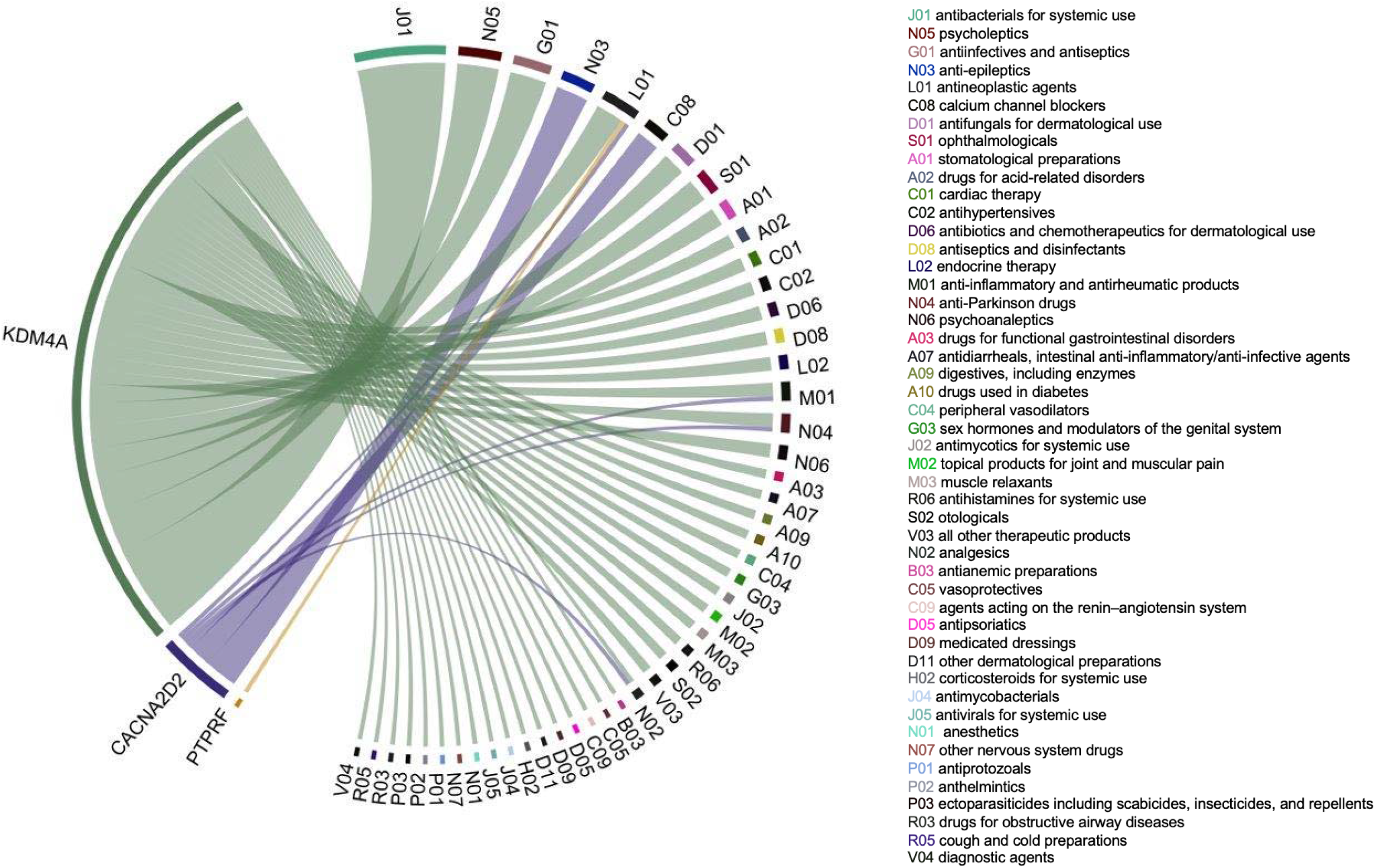
Chord diagram of genes significantly associated with POU at 10% FDR and the Anatomical Therapeutic Chemical classifications of drugs. The width of each line is determined by the number of drugs known to interact with each gene.

**Figure 5.**
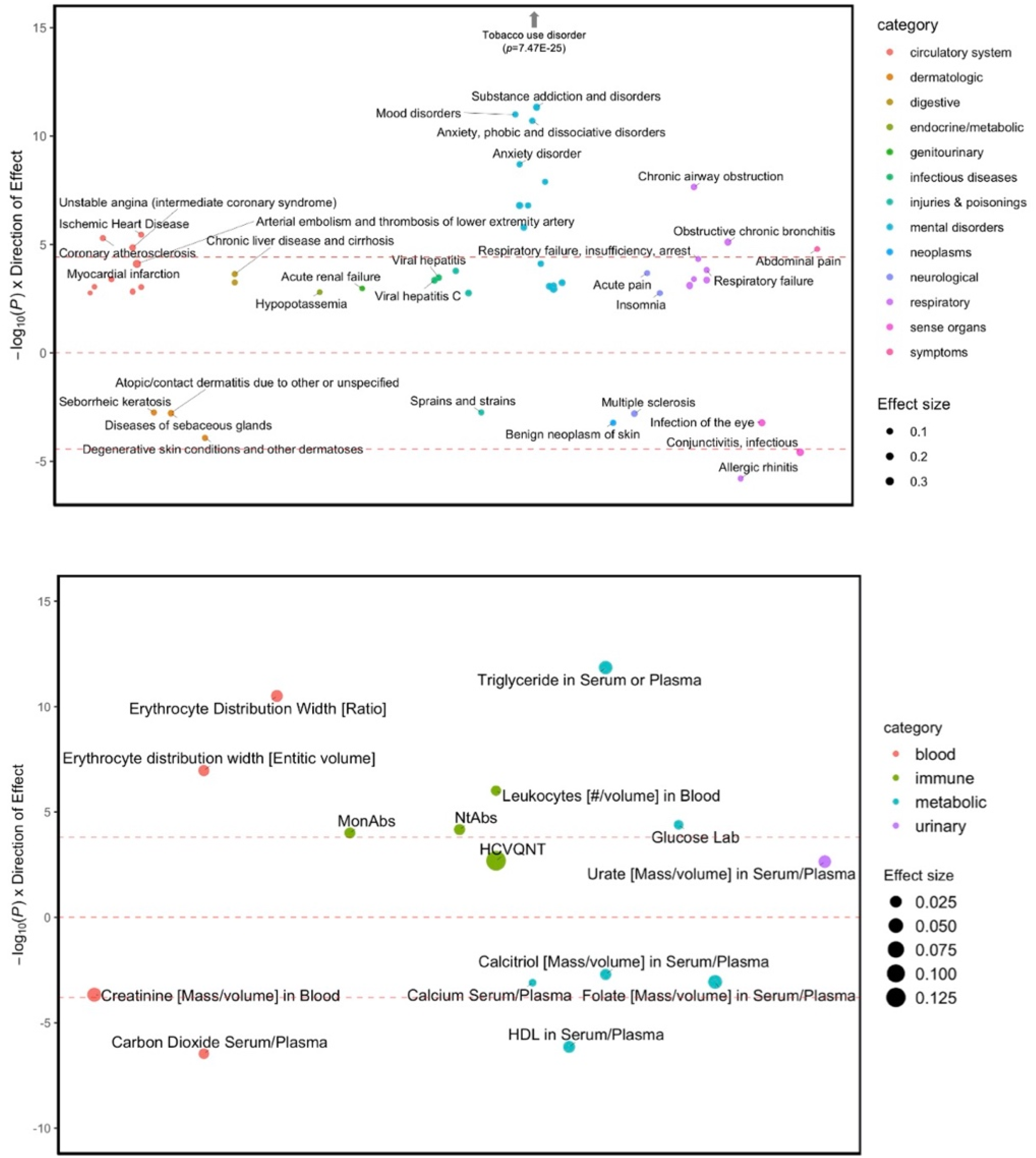
Phenome-wide (top) and lab-wide (bottom) association studies of polygenic risk scores for POU against 1,338 diseases and 315 biomarkers (respectively) available from BioVU. The top and bottom red dashed lines indicate the threshold for Bonferroni correction (PheWAS *p*=3.74E-05; LabWAS *p*=1.59E-04). Only FDR significant results are presented. The sizes of the dots correspond to the magnitude of the effect. Y axis represents the negative logarithm of the p-value for each trait multiplied by the sign of the effect.

### Genetic correlation analyses, and clustering solution

We performed a series of genetic correlations and identified consistent, moderate to strong associations with well-known OUD comorbid factors. We considered 935 traits, which represented 17 categories (substance use, psychiatric, impulsivity, personality, lifestyle, education, cognitive, health, longevity, metabolic, eyes, teeth, sleep, pain, reproductive, medication and anthropometric). We identified significant genetic associations between POU and 253 of these 935 traits (FDR 5%; **Supplementary Table 11**).

Notably, POU showed positive genetic correlations with OUD as measured by the Million Veteran Program (**MVP**, *r*_*g*_=0.64, *p*=9.97E-09, European), with opioid dependence versus unexposed controls (European) as measured by the Psychiatric Genomics Consortium (**PGC**, *r*_*g*_=0.80, *p*=3.24E-04; opioid dependence versus opioid exposed controls showed no significant heritability and therefore could not be tested), and with opioid medication use (*r*_*g*_=0.46, *p*=2.25E-12) in participants from UKB.

Beyond opioid-related traits, we also identified strong genetic correlations with alcohol, nicotine, and cannabis-related traits, from initiation (nicotine, *r*_*g*_=0.63, *p*=7.14E-38; cannabis, *r*_*g*_=0.35, *p*=1.06E-06), to high levels of consumption (drinks per week, *r*_*g*_=0.36; *p*=2.11E-15), to misuse (AUDIT-P: *r*_*g*_=0.42, *p*=5.60E-08) and dependence (alcohol, *r*_*g*_=0.74, *p*=7.21E-08; the Fagerström Test for Nicotine Dependence, *r*_*g*_=0.60, *p*=2.98E-10; cannabis use disorder, *r*_*g*_=0.63, *p*=2.36E-13), and other smoking behaviors, such as maternal smoking around birth (*r*_*g*_=0.64, *p*=3.44E-26). Interestingly, hierarchical HCA with AGNES found that POU clustered closely with substance use disorders, as opposed to consumption phenotypes (**Supplementary Figure 3**).

We also identified a number of associations with UKB pain phenotypes, including positive genetic correlations with several pain ICD diagnoses (*e*.*g*., pain in throat, pelvic pain, *r*_*g*_ range=[0.37-0.39], *p*<8.22E-05), and prevalent pain conditions such as back and knee pain, and headaches (*r*_*g*_ range=[0.21-0.42], *p*<8.50E-03). As expected, we found positive genetic associations with chronic pain (*r*_*g*_=0.42, *p*=4.04E-17), and higher pain relief medication intake (*e*.*g*., Ibuprofen, *r*_*g*_=0.49; *p*=1.61E-10).

We also identified positive genetic associations between POU and other behavioral and psychiatric traits, such as mood swings (*r*_*g*_=0.40, *p*=1.04E-13), risk-taking behavior (*r*_*g*_=0.38, *p*=2.09E-08), major depressive disorder (*r*_*g*_=0.44, *p*=1.59E-10), insomnia (*r*_*g*_=0.39, *p*=6.88E-11), loneliness (*r*_*g*_=0.28, *p*=4.81E-06) and irritability (*r*_*g*_=0.27, *p*=1.74E-05).

Lastly, POU was negatively correlated with educational attainment (*e*.*g*., obtaining a college or university degree, *r*_*g*_=-0.47; *p*=6.13E-25) and intelligence (*r*_*g*_=-0.41; *p*=7.22E-11).

These genetic correlations generally remained consistent after conditioning POU on risk-taking behavior, nicotine, and cannabis use disorders (**Supplementary Tables 12, 13** and **14** respectively). These associations were also broadly consistent when we conditioned on pain (**Supplementary Tables 16**), and OUD (**Supplementary Tables 17**). Intriguingly, when we corrected for alcohol dependence some of the associations increased, particularly with OUD and opioid dependence, cannabis use disorder, and depressive symptoms, whereas some associations dramatically decreased, such as nicotine dependence as measured via FTND (**Supplementary Table 15**) [43].

### Phenome-wide association analyses

We conducted PheWAS with EHR data to test the association between polygenic risk for POU and liability across thousands of medical conditions from hospital-based cohorts. Our PheWAS identified significant associations between POU and 53 medical traits across 13 categories (*i*.*e*., psychiatric disorders, respiratory, circulatory system, symptoms, sense organs, dermatologic, injuries and poisonings, neurological, digestive, infectious diseases, neoplasms, genitourinary, endocrine/metabolic; **Supplementary Table 18**). Similar to the genetic correlations, the strongest associations were with substance use disorders, including tobacco use disorder (OR=1.15, *p*=7.47E-25), alcohol-related disorders (OR=1.12, *p*=7.69E-05) and substance addiction and disorders (OR=1.17, *p*=4.67E-12), which included both opiates and other substances.

Another similarity between the PheWAS and the genetic correlations was the positive association between POU and pain phenotypes measured in EHR, such as acute pain (OR=1.05, *p*=2.09E-04) and chest pain (OR=1.04, *p*=8.90E-04). POU was also significantly associated with various psychiatric disorders, including mood disorders (OR=1.08, *p*=1.01E-11), anxiety disorders (OR=1.09, *p*=1.98E-11), depression (OR=1.07, *p*=1.29E-08) and posttraumatic stress disorder (OR=1.18, *p*=1.60E-07).

Finally, we discovered additional associations between POU and other EHR traits, including respiratory failure (OR=1.06, *p*=4.70E-05), insomnia (OR=1.06, *p*=1.75E-03) and ischemic heart disease (OR=1.06, *p*=3.57E-06).

We repeated the PheWAS analyses after adjusting for SUD and OUD diagnoses (**Supplementary Tables 19** and **20**). The magnitude of the effects persisted for most traits even though the strength of the association for a few conditions was diminished, particularly when controlling for general SUD.

### Lab-wide association analyses (LabWAS)

In addition to the PheWAS, we also examined laboratory results, which we refer to as LabWAS. The LabWAS identified 15 biomarkers significantly associated with POU (**Supplementary Table 21**). For instance, POU was associated with metabolic biomarkers, including increased triglyceride in serum or plasma (beta=0.04, *p*=1.41E-12) and blood glucose (beta=0.02, *p*=4.02E-5), and lower HDL cholesterol (beta=-0.03, *p*=7.21E-07), calcium (beta=-0.01, *p*=8.00E-04), folate (beta=-0.04, *p*=8.70E-04), and calcitriol (beta=-0.02, *p*=1.97E-03). We also found associations with blood-related biomarkers, such as high erythrocyte distribution (beta=0.03, *p*=3.14E-11), increased leukocytes (beta=0.02, *p*=9.68E-07), reduced carbon dioxide in serum/plasma (beta=-0.02, *p*=3.42E-7) and creatinine (beta=-0.04, *p*=2.15E-04).

Similar to the PheWAS results, the above-mentioned associations persisted after we corrected for SUD and OUD diagnoses (**Supplementary Tables 22** and **23**), suggesting the biomarkers are primarily affected by genetic liability for POU rather than substance use status.

## DISCUSSION

In this study, we performed a GWAS of problematic opioid use (‘ever taking prescription painkillers not as prescribed’) in 132,113 23andMe research participants of European ancestry. This represented a novel approach to studying OUD in a population-based cohort. Our results show that this single question captured a genetic signal that is correlated with signals from well-characterized cohorts that have been clinically diagnosed with OUD. Notably, the genetic correlations with OUD persisted even after correcting for risk-taking behavior and other putatively similar dimensional phenotypes. We also identified novel associations with laboratory-based biomarkers, demonstrating the overarching impact of POU on health. While previous GWAS of OUD have used clinically-ascertained cohorts, our results suggest that POU provides a cost-effective alternative to diagnosed OUD that is viable in non-clinically ascertained populations, making it possible to rapidly obtain large sample sizes that can aid in OUD gene discovery.

Obtaining a large enough sample size to effectively identify risk loci has been a common obstacle for OUD GWAS [44]. Our study represents a new and qualitatively different way of characterizing individuals at high risk for OUD. Our approach is stimulated by the idea of fractionating OUD and looking at POU as an early stage of misuse [45]. In particular, we were motivated to characterize the common mechanism of taking opioids not as prescribed, which can lead to abuse. Sometimes referred to as minimal phenotyping, where a complex trait is reduced to a single yes or no question [45], the polygenic signal of POU is nevertheless informative for aspects of OUD risk that is not intended to be a ‘noisy’ measurement of the true underlying disease.

The most important finding in this study is the identification of the polygenic architecture of POU that is highly comparable to the findings from GWAS of clinically-ascertained OUD cohorts. We observed a positive association of POU with OUD and opioid dependence measured by two of the largest available GWAS, MVP and PGC, and with a GWAS of opioid medication use in UKB participants. As might have been expected, we also observed strong positive genetic correlations with alcohol dependence and tobacco smoking, as well as with various psychiatric traits associated with OUD, including mood swings, risk-taking, anxiety, depression, and insomnia. These sets of genetic correlations mirror those from previous GWAS of clinically ascertained OUD samples [6–8]. However, we also showed that the overlap is not complete, and whether POU could be an early manifestation of risk for subsequent OUD is not directly explored by our study.

We initially speculated that some of the genetic signal associated with POU could be confounded by genetic factors unrelated to opioid misuse, such as risk-taking behaviors, comorbid substance use (such as tobacco or cannabis use disorders, and even OUD diagnosis) or pain. Previous reports have indicated that genetic studies using an unscreened sample for relevant comorbid conditions [6], including all individuals being exposed to opioids at least once, can introduce biases in the genetic analyses. However, when we adjusted for these known risk factors, the signal remained consistent, suggesting that we are capturing a signal that is specific to opioids and not merely confounded by signals associated with the secondary phenotypes.

This study has also revealed novel biological insights for opioid misuse. While we were unable to replicate the previously reported GWAS signals associated with OUD, we discovered two novel loci associated with POU. Rs640561 replicated in the largest available GWAS of OUD ([7]). Both lead SNPs (rs3791033, rs640561) and nearby genes have shown previous associations with other psychiatric traits, particularly smoking phenotypes, which are known to be highly comorbid with OUD and were also strongly correlated with POU. In particular, one of the strongest signals included variants in *KDM4A*, which is a protein coding gene that belongs to the Jumonji domain 2 (JMJD2) family of histone demethylases [46]. KDM4 demethylases catalyze the removal of the methyl marks (H3K9me3, H3K36me3), thereby regulating a range of crucial biological functions [47], including cell differentiation and proliferation. Intriguingly, the JMJD2 or KDM4 family have been shown to be important epigenetic regulators in cancer cells [48–50] and, relevant to this study, reward circuitry in depression [51] and alcohol withdrawal [52, 53]. The fact that *KDM4A* has known interactions with medications that are used to treat other psychiatric conditions, such as depression, or target dopaminergic or serotonergic systems, as well as drugs used in opioid anesthetics, suggests that KDM4 demethylases may be implicated in relevant biology for opioid behaviors and could represent promising druggable targets [54]. However, the biology of this locus could be more complex. Although the credible set analysis suggested that the most probable causal variants are located in *KDM4A*, the lead variant in this region, rs3791033, is an eQTL for several other nearby genes *PTPRF, HYI, ST3GAL3, MED8, CCDC24, ATP6V0B, WDR65, ARTN, TIE1*. Our second strongest signal was rs640561, which is near the gene *LRRIQ3*, which is a protein coding gene of leucine rich repeats and IQ Motif Containing 3. rs640561, or SNPs in strong LD, have been previously associated with other substance use traits, including smoking [55–57] and alcohol consumption [57], schizophrenia [58], education attainment and math ability [59], but the mechanism whereby this SNP affects OUD liability is largely unknown. Lastly, the gene-set analyses revealed that the polygenic signal for POU is implicated in pathways that modulate µ-opioid receptor sensitization. Taken together, these findings suggest a potential avenue for identifying new therapeutic targets for problematic opioid use.

We examined the genetic associations between POU PRS and thousands of clinical diagnoses and biomarkers measured in a hospital-based setting via PheWAS and LabWAS analyses. These analyses revealed significant associations for a wide range of health conditions, implicating POU across all major body systems, some of which had been previously associated with various clinical OUD cohorts and others that are novel.

For example, we consistently identified strong associations between POU and poor metabolic function (*e*.*g*., increased triglyceride levels, lower HDL cholesterol, high blood glucose levels), which can result in serious downstream health consequences. These results are consistent with previous studies showing poor metabolic function in individuals who misuse opioids [60], and in morphine-induced rats [61]. Some metabolic markers, such as calcium and folate, have been previously associated with opioid misuse [62, 63], whereas others (calcitriol) were identified as novel. Although it has been previously speculated that reduced metabolic biomarkers (calcium, folate) in high opioid users could be a direct consequence of repeated exposure to opioids, our results suggest that at least part of the previously reported correlations are due to common genetic factors that influence opiate use, calcium and folate. POU PRS was also associated with blood-related biomarkers, including increased erythrocyte and decreased lymphocyte counts, consistent with previous phenotypic studies [64, 65]. We observed an association between POU PRS and decreased carbon dioxide and creatinine levels, contrary to a previous finding showing that opioids can decrease sensitivity of peripheral chemoreceptors in the lung, leading to increased carbon dioxide levels [66], or no significant difference of creatinine levels among the heroin and opium participants studied [62], suggesting that some of these discrepancies may be associated with environmental consequences of OUD. It is also possible that some of the relationships that we identified could be a consequence of smoking and alcohol consumption, since high POU PRS was associated with tobacco and alcohol use disorders; whether the associations we report are a consequence of those relatively common behaviors is not directly tested in this study.

This study is not without limitations. The screening and composition of the control group is crucial for studies of substance use and abuse [6, 31, 44]. Our controls indicated they had never used opiates not as prescribed, which could include individuals who had simply never used an opiate, along with individuals who had anywhere from minimal to extensive experience with opiates but had never deviated from their prescribed use. POU might be even more useful if we had additional data that allowed us to exclude individuals that have never used opioids. Using an unscreened control group can lead to considerable phenotypic heterogeneity across samples [7, 67]. Similarly, it is unclear what type of prescription painkillers the subjects included in our study may have used. Future studies with improved phenotyping around this topic and greater sample size could be even more productive. A second important limitation is our inability to evaluate whether the individuals included in our analyses suffered from mild versus severe pain. Although high genetic predisposition for chronic pain may itself be a risk factor for OUD, this concern was partially addressed by our mtCOJO analysis in which we conditioned on pain. That analysis revealed that the association between POU and OUD persevered even after correcting for pain, suggesting that POU was not correlated with OUD via its ability to capture genetic predisposition to pain. Lastly, the 23andme participant base was not ascertained for OUD and is more educated and has a higher socioeconomic status than the broader population; therefore, it is possible that a similar misuse phenotype captured in other higher risk populations may yield different results.

In summary, we have shown that the genetic signature for opioid misuse that we were able to capture via self-report in an unselected population is similar to genetic risk for OUD. Our work sets the stage for future analyses incorporating a multivariate framework (*e*.*g*., genomic Structural Equation Modelling), and larger sample sizes. Our approach provides new, cost-efficient tools for genetic research related to OUD and provides insights into an intermediate behavior that may set the stage for later transition to OUD.

## Data Availability

We have provided summary statistics for the top 10,000 SNPs (Supplementary Table 24). Full GWAS summary statistics will be made available through 23andMe to qualified researchers under an agreement with 23andMe that protects the privacy of the 23andMe participants. Interested investigators should and reference this paper for more information.

## Data availability

We have provided summary statistics for the top 10,000 SNPs (**Supplementary Table 24**). Full GWAS summary statistics will be made available through 23andMe to qualified researchers under an agreement with 23andMe that protects the privacy of the 23andMe participants. Interested investigators should email dataset- and reference this paper for more information.

## Acknowledgements

We would like to thank the research participants and employees of 23andMe (Michelle Agee, Babak Alipanahi, Adam Auton, Robert K. Bell, Katarzyna Bryc, Sarah L. Elson, Pierre Fontanillas, Nicholas A. Furlotte, David A. Hinds, Karen E. Huber, Aaron Kleinman, Nadia K. Litterman, Jennifer C. McCreight, Matthew H. McIntyre, Joanna L. Mountain, Elizabeth S. Noblin, Carrie A.M. Northover, Steven J. Pitts, J. Fah Sathirapongsasuti, Olga V. Sazonova, Janie F. Shelton, Suyash Shringarpure, Chao Tian, Joyce Y. Tung, Vladimir Vacic, and Catherine H. Wilson) for making this work possible.

We would also like to thank The Externalizing Consortium, for sharing the GWAS summary statistics of externalizing. The Externalizing Consortium: Principal Investigators: Danielle M. Dick, Philipp Koellinger, K. Paige Harden, Abraham A. Palmer. Lead Analysts: Richard Karlsson Linnér, Travis T. Mallard, Peter B. Barr, Sandra Sanchez-Roige. Significant Contributors: Irwin D. Waldman. The Externalizing Consortium has been supported by the National Institute on Alcohol Abuse and Alcoholism (R01AA015416 -administrative supplement), and the National Institute on Drug Abuse (R01DA050721). Additional funding for investigator effort has been provided by K02AA018755, U10AA008401, P50AA022537, as well as a European Research Council Consolidator Grant (647648 EdGe to Koellinger). The content is solely the responsibility of the authors and does not necessarily represent the official views of the above funding bodies. The Externalizing Consortium would like to thank the following groups for making the research possible: 23andMe, Add Health, Vanderbilt University Medical Center’s BioVU, Collaborative Study on the Genetics of Alcoholism (COGA), the Psychiatric Genomics Consortium’s Substance Use Disorders working group, UK10K Consortium, UK Biobank, and Philadelphia Neurodevelopmental Cohort.

