## Supplementary Material for "Genome-wide association study of problematic opioid prescription use in 132,113 23andMe research participants of European ancestry"

**Supplementary Methods**

**Genotyping, imputation and quality control**

Samples were genotyped on a 23andMe custom genotyping array platform (Illumina HumanHap550+ Bead chip V1 V2, OmniExpress+ Bead chip V3, Custom array V4, and Infinium Global Screening Array V5).  Samples had minimum call rates of 98.5%. A total of 1,609,130 SNPs and InDels (Insertion/Deletion) were genotyped across all platforms (**Supplementary Table 1**; see [[1–7]](https://www.zotero.org/google-docs/?eNxePg) for extended genotyping and sample quality details).

A maximal set of unrelated individuals was chosen for the analysis using a segmental identity-by-descent (**IBD**) estimation algorithm [[4]](https://www.zotero.org/google-docs/?v9OP76) to ensure that only unrelated individuals were included in the sample. Individuals were defined as related if they shared more than 700 cM IBD, including regions where the two individuals shared either one or both genomic segments IBD. This level of relatedness (~20% of the genome) corresponds to approximately the minimal expected sharing between first cousins in an outbred population.

We imputed participant genotype data against a reference panel combining the September 2013 release of 1000 Genomes phase 3 reference haplotypes imputation reference panel [[8]](https://www.zotero.org/google-docs/?zattSm). We phased and imputed data for each genotyping platform separately. We phased using an internally developed phasing tool, Finch, which implements the Beagle haplotype graph-based phasing algorithm [[5]](https://www.zotero.org/google-docs/?5urMvg), modified to separate the haplotype graph construction and phasing steps. In preparation for imputation, we split phased chromosomes into segments of no more than 10,000 genotyped SNPs, with overlaps of 200 SNPs. We excluded SNPs with Hardy-Weinberg equilibrium *p <* 10^−20^, call rate < 95%, or with large allele frequency discrepancies compared to European 1000 Genomes reference data. Frequency discrepancies were identified by computing a 2 x 2 table of allele counts for European 1000 Genomes samples and 2000 randomly sampled 23andMe customers with European ancestry, and identifying SNPs with a 𝜒^2^ *p* < 10^−15^. We imputed each phased segment against all-ethnicity 1000 Genomes haplotypes (excluding monomorphic and singleton sites) using Minimac3 [[6]](https://www.zotero.org/google-docs/?1KZZiD), using 5 rounds and 200 states for parameter estimation. After quality control, we analyzed 11,311,983 SNPs.

For the X chromosome, we built separate haplotype graphs for the non-pseudoautosomal region and each pseudoautosomal region, and these regions were phased separately. We then imputed males and females together using Minimac3, as with the autosomes, treating males as homozygous pseudo-diploids for the non-pseudoautosomal region.

For tests using imputed data, we use the imputed dosages rather than best-guess genotypes. We imputed HLA allele dosages from SNP genotype data using HIBAG [[7]](https://www.zotero.org/google-docs/?jhm9Co). We imputed alleles for HLA-A, B, C, DPB1, DQA1, DQB1, and DRB1 loci at four-digit resolution. To test associations between HLA allele dosages and phenotypes, we performed linear regression using the same set of covariates used in the SNP-based GWAS. We performed separate association tests for each imputed allele. HLA analysis did not identify any significant signal and hence is not described in the main text.

**Supplementary Figures**

**Supplementary Figure 1.** Regional association plot focusing on top SNP rs3791033 on chromosome 1 at position 44.1 Mb. The regional association plot shows association test statistics versus position in the vicinity of the strongest associations. The -log10(P value) is shown on the left y-axis; position in Mb is on the x-axis. Recombination rates (expressed in centiMorgans cM per Mb; NCBI Build GRCh37; highlighted in blue) are shown on the right y-axis. Pairwise linkage disequilibrium (r2) of each SNP with the top SNP in the region is indicated by its color. A ‘+’ indicates an imputed variant, an ‘x’ indicates an imputed protein-altering variant, an ‘o’ symbol indicates a genotyped variant. We also report details of the credible set, calculated under the assumption of a single causal variant within the locus.

**Supplementary Figure 2.** Regional association plot focusing on top SNP rs640561 on chromosome 1 at position 73.9 Mb. The regional association plot shows association test statistics versus position in the vicinity of the strongest associations. The -log10(P value) is shown on the left y-axis; position in Mb is on the x-axis. Recombination rates (expressed in centiMorgans cM per Mb; NCBI Build GRCh37; highlighted in blue) are shown on the right y-axis. Pairwise linkage disequilibrium (r2) of each SNP with the top SNP in the region is indicated by its color. A ‘+’ indicates an imputed variant, an ‘x’ indicates an imputed protein-altering variant, an ‘o’ symbol indicates a genotyped variant. We also report details of the credible set, calculated under the assumption of a single causal variant within the locus.

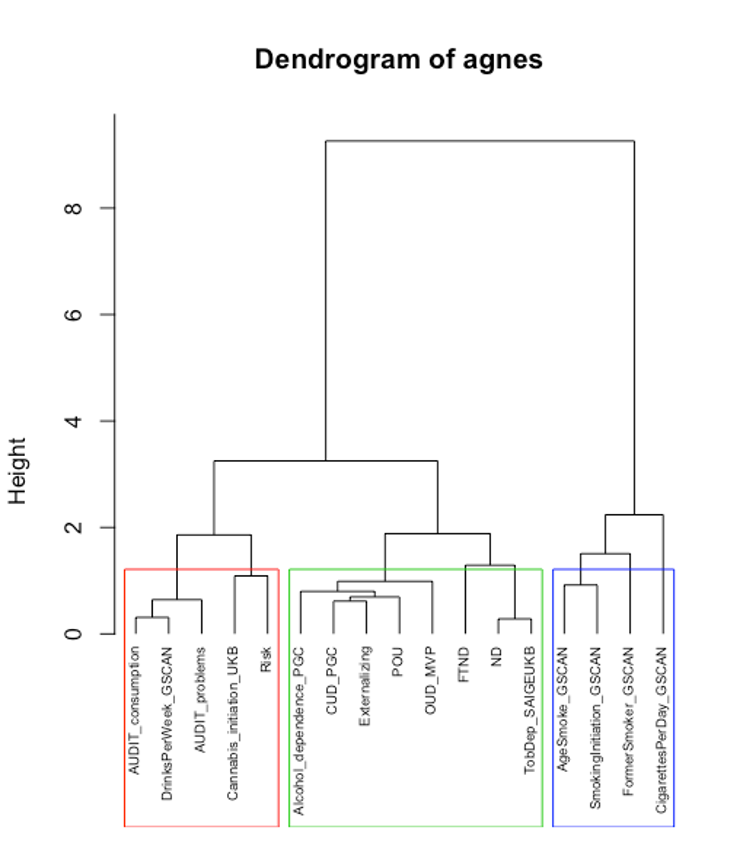

**Supplementary Figure 3. Hierarchical Clustering Analysis with Agglomerative Nesting (HCA with AGNES) dendrogram.** Input was the pairwise genetic correlation matrix. Boxes represent the division in branches needed to divide substance use and use disorder traits. Height (y-axis) represents distance metrics between the clusters. Interpreting from the bottom-up, POU clustered on its own. The next level of associations was with cannabis use disorder and externalizing psychopathology; these traits clustered with alcohol dependence, and next opioid use disorder broadly, before the branch converged with measures of nicotine dependence. Interpreting from the top-down, we noticed two clusters, a nicotine behaviors cluster and a cluster of all other substance use behaviors. One more branch was needed to gain specificity between use and use disorder clusters, which formed three branches. The middle branch was as substance use disorder branch with clustered POU with use disorders.
